# An immune-protein signature combining TRAIL, IP-10 and CRP for accurate prediction of severe COVID-19 outcome

**DOI:** 10.1101/2021.06.27.21259196

**Authors:** Niv Samuel Mastboim, Alon Angel, Oded Shaham, Tahel Ilan Ber, Roy Navon, Einav Simon, Michal Rosenberg, Yael Israeli, Mary Hainrichson, Noa Avni, Eran Reiner, Paul Feigin, Kfir Oved, Boaz Tadmor, Pierre Singer, Ilya Kagan, Shaul Lev, Dror Diker, Amir Jarjou’i, Ramzi Kurd, Eli Ben-Chetrit, Guy Danziger, Cihan Papan, Sergey Motov, Ma’anit Shapira, Michal Stein, Adi Klein, Tanya M Gottlieb, Eran Eden

## Abstract

**BACKGROUND:** Accurately identifying COVID-19 patients at-risk to deteriorate remains challenging. Tools integrating host-protein expression have proven useful in determining infection etiology and hold potential for prognosticating disease severity.

**METHODS:** Adults with COVID-19 were recruited at medical centers in Israel, Germany, and the United States. Severe outcome was defined as intensive care unit admission, non-invasive or invasive ventilation, or death. Tumor necrosis factor related apoptosis inducing ligand (TRAIL) and interferon gamma inducible protein-10 (IP-10; also known as CXCL10) and C-reactive protein (CRP) were measured using an analyzer providing values within 15 minutes. A signature indicating the likelihood of severe outcome was derived generating a score (0-100). Patients were assigned to 4 score bins.

**RESULTS:** Between March and November 2020, 518 COVID-19 patients were enrolled, of whom 394 were eligible, 29% meeting a severe outcome. The signature’s area under the receiver operating characteristic curve (AUC) was 0.86 (95% confidence interval: 0.81-0.91). Performance was not confounded by age, sex, or comorbidities and superior to IL-6 (AUC 0.77; p = 0.033) and CRP (AUC 0.78; p < 0.001). Likelihood of severe outcome increased significantly (p < 0.001) with higher scores. The signature differentiated patients who further deteriorated after meeting a severe outcome from those who improved (p = 0.004) and projected 14-day survival probabilities (p < 0.001).

**CONCLUSION:** The derived immune-protein signature combined with a rapid measurement platform is an accurate predictive tool for early detection of COVID-19 patients at-risk for severe outcome, facilitating timely care escalation and de-escalation and appropriate resource allocation.

**FUNDING:** MeMed funded the study

## INTRODUCTION

Globally as of May 2021, there have been 155 million confirmed cases of severe acute respiratory syndrome coronavirus 2 (SARS-CoV-2) infection and almost 3.3 million deaths attributed to the resulting coronavirus disease 2019 (COVID-19) (https://coronavirus.jhu.edu/map.html). With vaccination campaigns taking time to implement, hospitals around the globe are still dealing with COVID-19 patients. Moreover, global herd immunity seems improbable given vaccination inequity and hesitancy combined with the emergence of variants (1, 2). Accordingly, it is predicted that SARS-CoV-2 will become endemic (3).

An unusual feature of SARS-CoV-2 is that it causes a broad spectrum of disease severity, ranging from asymptomatic infection to critical illness. While the vast majority of patients with SARS-CoV-2 infection develop mild to moderate disease (https://www.cdc.gov/coronavirus/2019-ncov/hcp/clinical-guidance-management-patients.html), without requiring supplemental oxygen therapy or hospitalization, up to 20% of patients experience life-threatening disease (4). The acute phase of SARS-CoV-2 infection is divided into two stages that correlate with the clinical manifestations and severity of the disease (5–7). The first acute stage is characterized by viral replication and relatively mild symptoms, while the second is characterized by a dysregulated immune response that can rapidly progress to severe symptoms and critical complications including respiratory failure and death (8–13). Patient management is based on identifying the pathophysiological stage and directing appropriate interventions as well as accurate assessment of the likelihood that a patient will progress from one stage to another. Despite major advances in COVID-19 patient management, predicting which patients have increased risk for severe outcome remains challenging. Early prediction in COVID-19 is of particular importance because high-risk patients may benefit from earlier escalation of care (i.e., intensive care unit (ICU) placement, immunomodulation treatment) (3), whereas low-risk patients may benefit from avoiding unnecessary treatments or hospital admission. Accurate risk stratification is also helpful when assessing response to treatment and for discharge decisions.

Current clinical scores that incorporate patient demographics (e.g., age, sex), vital signs, comorbidities and radiographic images, are either indicative of organ damage that has already occurred (i.e., not predictive), fail to capture the complexity of COVID-19 disease (i.e., not accurate) or are difficult to integrate in routine decision making (i.e., not practical) (14). Multiple studies indicate that individual markers, including Interleukin-6 (IL-6), procalcitonin (PCT), ferritin, D-dimer, creatinine, CD3 and CD4 T-cell counts are associated with severe COVID-19 disease outcome (13, 15, 16). However, the utility of individual biomarkers is typically constrained by uncertainty regarding result interpretation or poor potential to identify severe disease at early stages (17). Further, combining these markers is either impractical (difficult to measure, time consuming or require multiple technologies) or has not proven to significantly increase predictive performance as the composite parameters provide overlapping information and are not viral-specific (14). Given the complexity of the immunopathogenesis of COVID-19, a signature combining several markers from distinct biological pathways is more likely to serve as a broadly applicable and accurate tool (15, 16). The number of biomarkers should be practicable and measurable so the tool can be clinically useful.

Recently, a platform was developed that measures in fifteen minutes two viral-induced biomarkers - tumor necrosis factor related apoptosis inducing ligand (TRAIL) and interferon gamma inducible protein-10 (IP-10; also known as CXCL10) – as well as the inflammatory biomarker, C-reactive protein (CRP). TRAIL regulates immune responses via apoptosis, with lower levels associated with severe infection, including sepsis (18–21). TRAIL levels increase in viral infection (22–24) and are reduced in severe bacterial and viral infections, making it a particularly useful biomarker for severe viral infection. In COVID-19 disease, low TRAIL levels are associated with inability to clear the virus and disease severity (25, 26). IP-10 expression is also induced by viral infection (22–24). A regulator of inflammatory and endothelial cells, IP-10 has been implicated as a player in the lung injury during dysregulated responses to severe viral infection (27). Accumulating evidence identifies IP-10 as an early marker of COVID-19 progression, with maintained high levels associated with mortality (15, 25, 26, 28, 29). Induced expression of CRP, a regulator of inflammation (30), is typically observed in response to bacterial infection (31, 32) but also found to be associated with severe SARS-CoV-2 infection (33–35). As each one of these biomarkers captures a different dimension of disease progression to severe outcome, their computational integration has the potential to produce a high performing severity score. Taken together, the severity score along with the rapid measurement platform, have potential to serve as an accurate and actionable test for early indication of COVID-19 disease progression.

This study reports derivation of an algorithm that computationally integrates the levels of TRAIL, IP-10, and CRP into a single numeric score that is an early indicator of severe outcome for COVID-19 patients (‘severity signature’). Such a signature can identify patients likely to deteriorate or not and in this way, help with objective decision making as to who may benefit from escalation or de-escalation of care as well as targeted (personalized) therapies (36) (https://www.covid19treatmentguidelines.nih.gov/critical-care/oxygenation-and-ventilation/).

## RESULTS

### Study population

Between March and November 2020, 518 Israeli, German, and USA SARS-CoV-2 positive patients were enrolled across six participating medical centers (Supplementary Table 1). This period encompassed the first and second COVID-19 waves in Israel, the first wave in Germany and the first and second waves in the US (Supplementary Figure 1). After exclusion of 124 patients that failed to meet eligibility criteria, the resulting derivation cohort comprised 394 patients (Figure 1). A composite severity outcome was defined based on mortality or respiratory failure requiring ICU level of care. 113 (29%) patients met the composite severity outcome, of whom 30 patients (27%) died.

**Figure 1.**
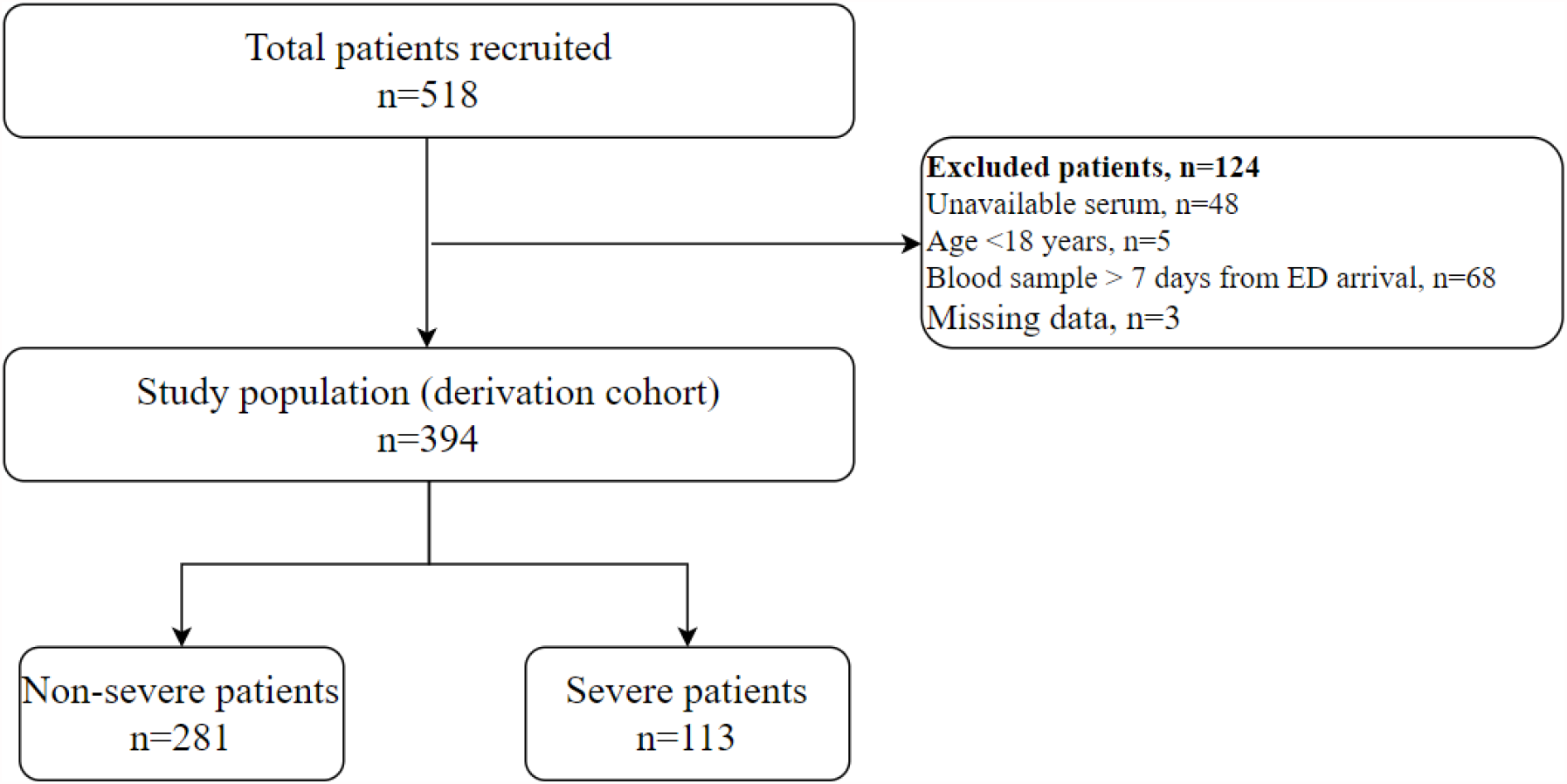
Derivation cohort

Age ranged from 19 to 98 years, with 59.1% male (Table 1). Patients who met severe outcomes were older, more likely to be male, and exhibited abnormal vital signs and elevated inflammatory markers on the day of blood draw as compared to non-severe patients. 18.8% of cohort patients (74/394) were hospitalized for a short duration (1-2 days) and did not meet a severe outcome.

**Table 1.** Clinical characteristics of the derivation cohort. IQR, interquartile range.

|  | Statistics | Study population<br>(n=394) | Severe, S<br>(n=113) | Non-Severe, NS (n=281) | S vs NS<br>p-value |
| --- | --- | --- | --- | --- | --- |
| Age (years) | median (IQR) | 61.5 (25.8) | 65.0 (21.0) | 60.0 (27.0) | 0.009 |
| Male sex | n (%) | 233 (59.1%) | 80 (70.8%) | 153 (54.4%) | 0.003 |
| Time from ED arrival to blood draw (days) | median (IQR) | 1.0 (2.0) | 2.0 (2.0) | 1.0 (2.0) | <0.001 |
| Length of hospital stay (days)<sup>A</sup> | median (IQR) | 6.5 (10.0) | 11.0 (13.8) | 5.0 (8.0) | <0.001 |
| Short hospital stay (1-2 days)<sup>A</sup> | n (%) | 76 (19.9%) | 2 (1.9%) | 74 (26.8%) | <0.001 |
| **Outcome<sup>B</sup>** |  |  |  |  |  |
| ICU admission | n (%) | 51 (12.9%) | 51 (45.1%) | 0 (0.0%) | N/A |
| Non-invasive ventilation | n (%) | 65 (16.5%) | 65 (57.5%) | 0 (0.0%) | N/A |
| Intubation with mechanical ventilation | n (%) | 49 (12.4%) | 49 (43.4%) | 0 (0.0%) | N/A |
| Mortality | n (%) | 30 (7.6%) | 30 (26.5%) | 0 (0.0%) | N/A |
| **Comorbidities** |  |  |  |  |  |
| Hypertension | n (%) | 163 (41.4%) | 54 (47.8%) | 109 (38.8%) | 0.114 |
| Diabetes mellitus | n (%) | 114 (28.9%) | 39 (34.5%) | 75 (26.7%) | 0.140 |
| Chronic heart failure | n (%) | 18 (4.6%) | 7 (6.2%) | 11 (3.9%) | 0.423 |
| Malignancy | n (%) | 19 (4.8%) | 10 (8.8%) | 9 (3.2%) | 0.034 |
| **Vital signs<sup>A,C</sup>** |  |  |  |  |  |
| Heart rate > 100 beats per minute | n (%) | 51 (17.3%) | 16 (18.4%) | 35 (16.9%) | 0.739 |
| Temperature ≥ 37.8°C | n (%) | 45 (18.1%) | 19 (28.4%) | 26 (14.4%) | 0.015 |
| Systolic blood pressure < 90 mmHg | n (%) | 8 (3.0%) | 6 (7.5%) | 2 (1.1%) | 0.010 |
| Respiratory rate > 20 breath/minute | n (%) | 32 (29.6%) | 20 (44.4%) | 12 (19.0%) | 0.006 |
| **Oxygen saturation on room air**<br>**(SpO<sub>2</sub>) ≤ 93%<sup>D</sup>** | **n (%)** | 91 (36.3%) | 32 (54.2%) | 59 (30.7%) | 0.002 |
| **Biomarkers<sup>A,C</sup>** |  |  |  |  |  |
| **WBC (K/ml<sup>3</sup>)** | **median (IQR)** | 6.4 (3.3) | 7.8 (3.2) | 5.8 (3.0) | <0.001 |
| **Ferritin (ng/ml)** | **median (IQR)** | 438.4 (1281.4) | 888.8<br>(1237.7) | 392.2 (864.2) | 0.026 |
| **CRP (mg/L)** | **median (IQR)** | 66.0 (139.8) | 166.8<br>(178.0) | 42.4 (83.7) | <0.001 |
| **PCT (ng/ml)** | **median (IQR)** | 0.1 (0.2) | 0.3 (0.5) | 0.1 (0.1) | <0.001 |
| **IL-6 (pg/ml)** | **median (IQR)** | 19.2 (44.3) | 50.9 (113.6) | 14.5 (34.6) | <0.001 |

<sup>A</sup>Missing data for a subset of cohort patients (detailed in Methods and Supplementary Table 11).
<sup>B</sup>Outcomes met before or within 14 days of blood draw.
<sup>C</sup>Measured on day of blood draw.
<sup>D</sup>https://www.covid19treatmentguidelines.nih.gov/

### A signature that integrates TRAIL, IP-10 and CRP accurately stratifies COVID-19 patients by disease severity

The host proteins TRAIL, IP-10 and CRP were differentially expressed in sera of severe versus non-severe COVID-19 patients (Figure *2*). Patients meeting severe outcomes exhibited significantly higher levels of CRP (median 167 vs. 42 mg/L; p < 0.001) and IP-10 (median 1632 vs. 420 pg/ml: p < 0.001) and lower levels of TRAIL (median 31 vs. 61 pg/ml; p < 0.001).

A signature integrating the serum concentrations of the three proteins was developed using the entire derivation cohort (see Methods). The signature score ranges from 0 to 100, with higher levels reflecting a higher likelihood for severe outcome. Signature scores were significantly higher in severe as compared to non-severe patients (median 78 vs. 26 score units; p < 0.001; Figure 3). The signature performance as indicated by area under the receiver operating characteristic curve (AUC) was 0.86 (95% CI: 0.81-0.91).

**Figure 2.**
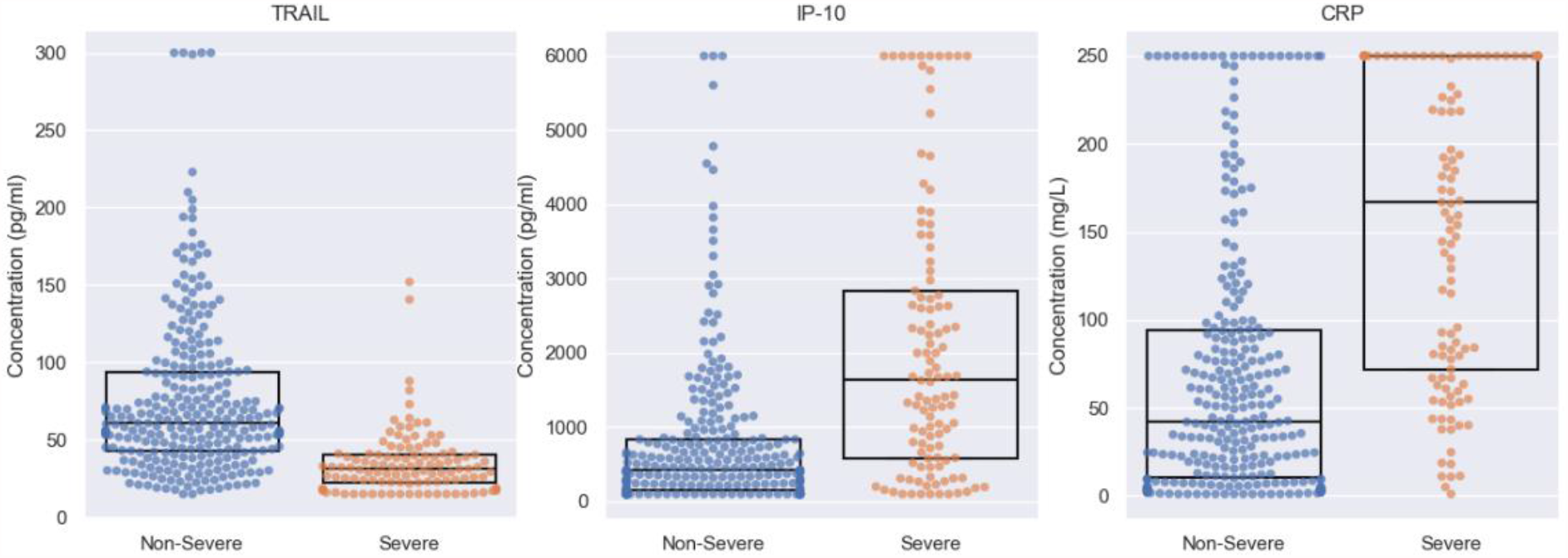
Differential expression of TRAIL, IP-10 and CRP in severe and non-severe COVID-19 infection. Dots represent patients and boxes denote median and interquartile range (IQR).

**Figure 3.**
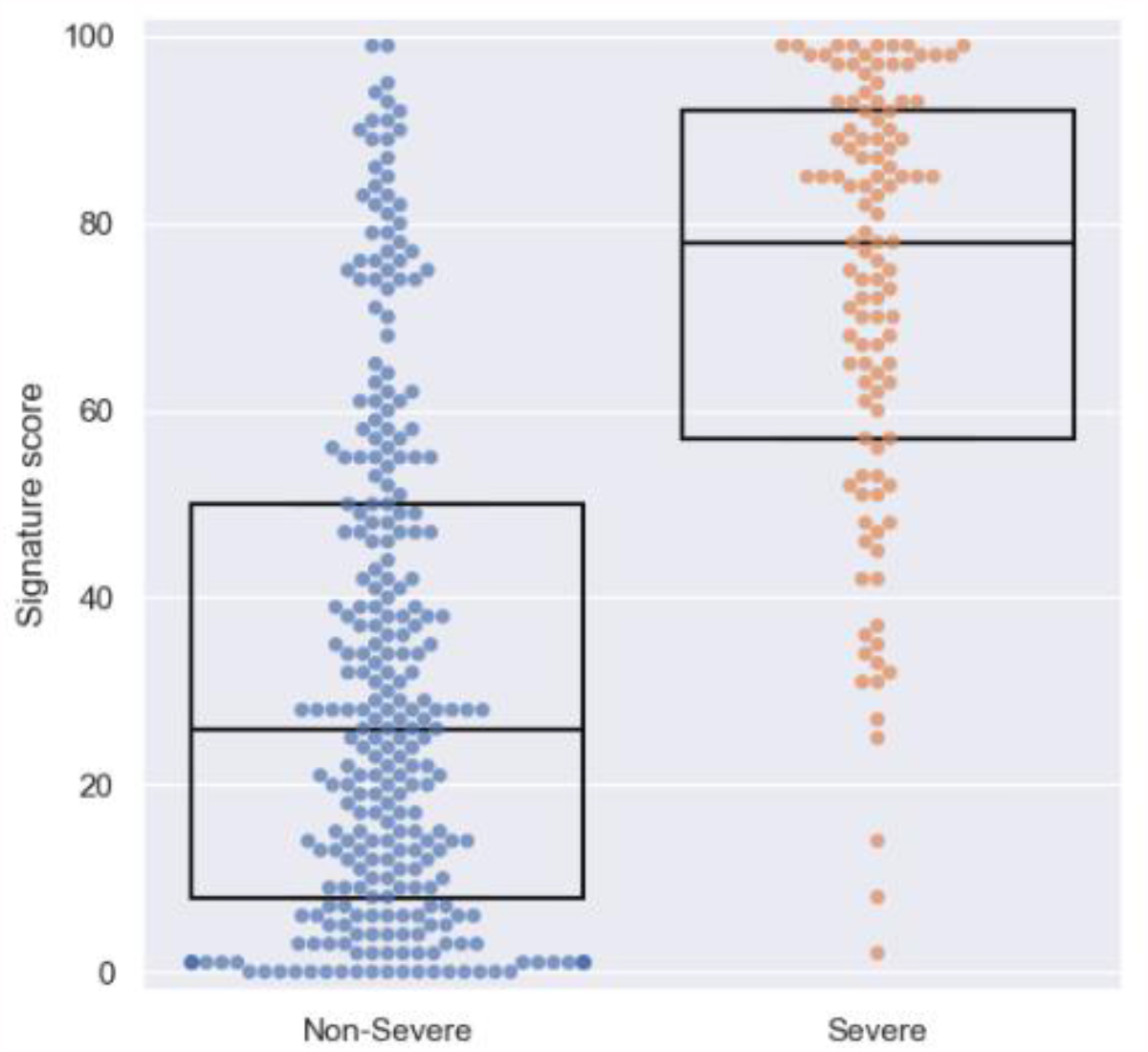
Signature score in severe and non-severe COVID-19 infection. Dots represent patients and boxes denote median and IQR.

To assess the signature’s potential for generalization its performance was compared to that of cross-validation based on the same cohort of patients (see Methods) (37). Cross-validation AUC was 0.86 (95% CI: 0.81-0.90), indistinguishable from the signature score, indicating the model was not overfitted and is generalizable.

To generate a clinically intuitive tool for severity stratification, four score bins were defined. Each patient was assigned to a bin based on their signature score, and within the bin according to their severity outcome. In this framework, the signature’s performance was demonstrated by a significant increase of the likelihood of COVID-19 severe outcome across the four bins (Cochran-Armitage, CA p < 0.001;Table 2). Only 3 severe patients (3% of all severe patients) were assigned to the lowest bin; clinical details for these patients are given in Supplementary Table 2. The proportion of patients intubated with mechanical ventilation or died increased across the four bins (Supplementary Table 3).

**Table 2.** Distribution of patients across score bins.

| *Bin* | *Score* | *n Total* | *%* | *n* | *n Non-* | *% Severe* | *% Non-Severe* | *LR (95% CI)* |
| --- | --- | --- | --- | --- | --- | --- | --- | --- |
|  |  |  | *Patients* | *Severe* | *Severe* | *(PPV)* | *(NPV)* |  |
| **4** | **80 < score ≤ 100** | 75 | 19.04 | 54 | 21 | 72.00 | 28.00 | 6.39 (4.06-10.07) |
| **3** | **40 ≤ score ≤ 80** | 116 | 29.44 | 46 | 70 | 39.66 | 60.34 | 1.63 (1.21-2.21) |
| **2** | **20 < score < 40** | 79 | 20.05 | 10 | 69 | 12.66 | 87.34 | 0.36 (0.19-0.67) |
| **1** | **0 ≤ score ≤ 20** | 124 | 31.47 | 3 | 121 | 2.42 | 97.58 | 0.06 (0.02-0.19) |
|  | **Total** | 394 | 100.00 | 113 | 281 |  |  |  |

LR, likelihood ratio. PPV, positive predictive value. NPV, negative predictive value.

### Signature maintained high performance across different sub-populations

To test whether the relationship between signature score and severe outcome is confounded by known risk factors, performance was inspected across sub-populations. Potential confounders evaluated included sex, age, and number of comorbidities. The time from emergency department (ED) arrival to blood draw was also evaluated as a surrogate for time from symptom onset. A significant increase in the likelihood of severe outcome with increasing signature score was observed in each of the sub-populations tested (Supplementary Table 4; p < 0.001 for each of the eight sub-populations). Furthermore, adding each of these risk factors to the host proteins did not improve the performance (Supplementary Table 5), indicating that the information contained in sex, age, number of co-morbidities and time from ED arrival is already captured by the status of the immune response.

### The signature outperforms known risk factors and candidate severity tools

Several parameters (e.g., age, oxygen saturation) (38) and biomarkers (e.g., IL-6) (33) have been proposed as candidate tools to aid the clinician in COVID-19 severity stratification. The signature AUC compared favorably with various biomarkers and parameters (Figure 4). The signature AUC was higher also when the comparison analysis was restricted to the subset of patients with available measurements for each comparator (Table 4), and when the definition of severe outcome was limited to intubation with mechanical ventilation or death (Supplementary Table 6).

**Figure 4.**
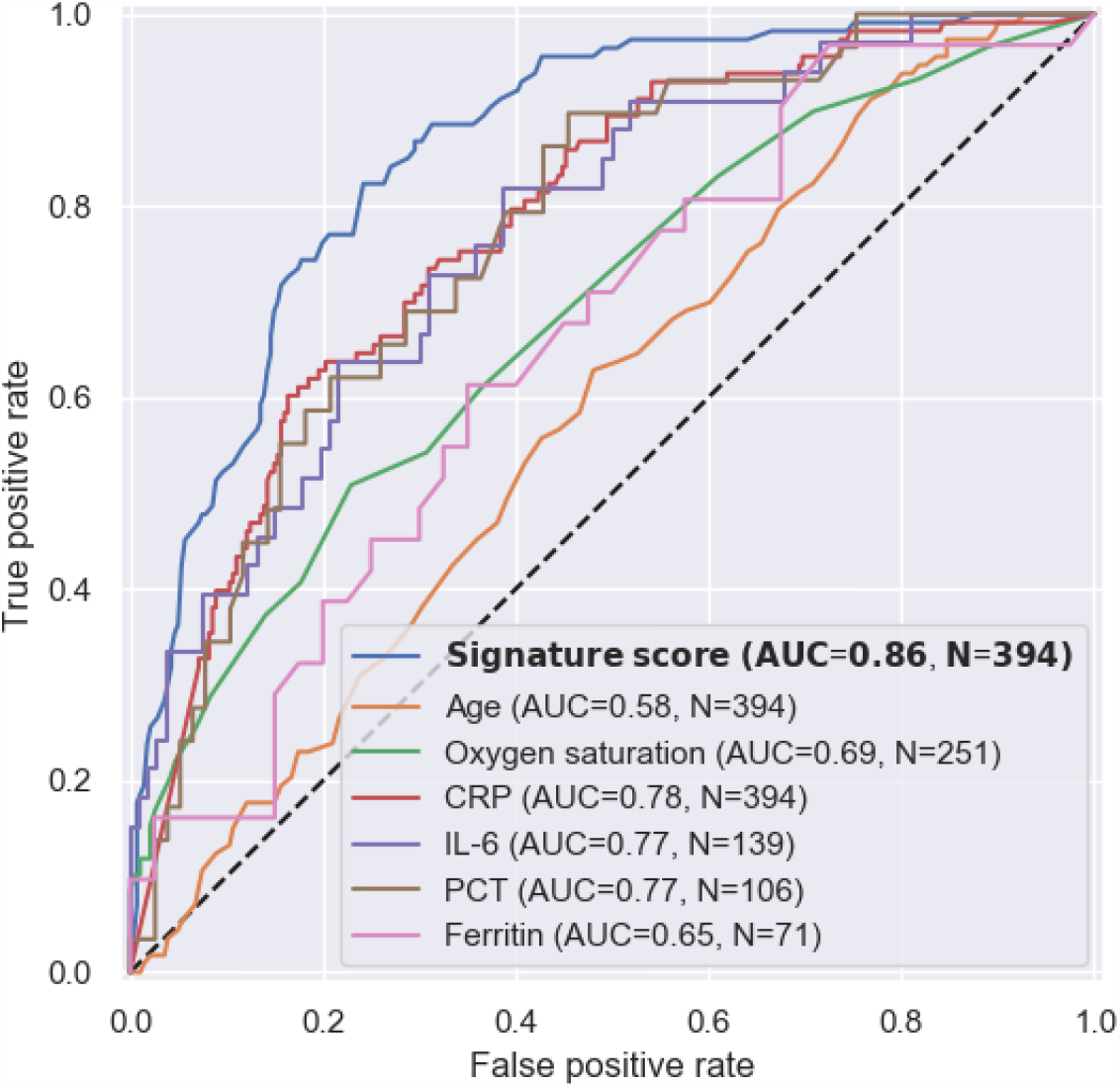
Receiver operating characteristic (ROC) curve of the signature compared to known risk factors and candidate severity tools.

**Table 3.** Comparison of the signature performance to other parameters and biomarkers on the subset of patients with available measurements for each comparator.

|  | **Total**<br>**available** | **n**<br>**Severe** | **n Non-**<br>**severe** | **Comparator**<br>**AUC (95% CI)** | **Signature**<br>**AUC (95% CI)** | **p-value** |
| --- | --- | --- | --- | --- | --- | --- |
| **Signature** | 394 | 113 | 281 | N/A | 0.86 (0.81-0.91) | N/A |
| **Age** | 394 | 113 | 281 | 0.58 (0.52-0.65) | 0.86 (0.81-0.91) | <0.001 |
| **Oxygen**<br>**saturation** | 251 | 59 | 192 | 0.69 (0.60-0.77) | 0.89 (0.84-0.95) | <0.001 |
| **CRP** | 394 | 113 | 281 | 0.78 (0.73-0.83) | 0.86 (0.81-0.91) | <0.001 |
| **IL-6** | 139 | 33 | 106 | 0.77 (0.67-0.87) | 0.89 (0.81-0.97) | 0.033 |
| **PCT** | 106 | 29 | 77 | 0.77 (0.66-0.88) | 0.90 (0.82-0.98) | 0.039 |
| **Ferritin** | 71 | 31 | 40 | 0.65 (0.52-0.78) | 0.80 (0.69-0.91) | 0.054 |

**Table 4.**
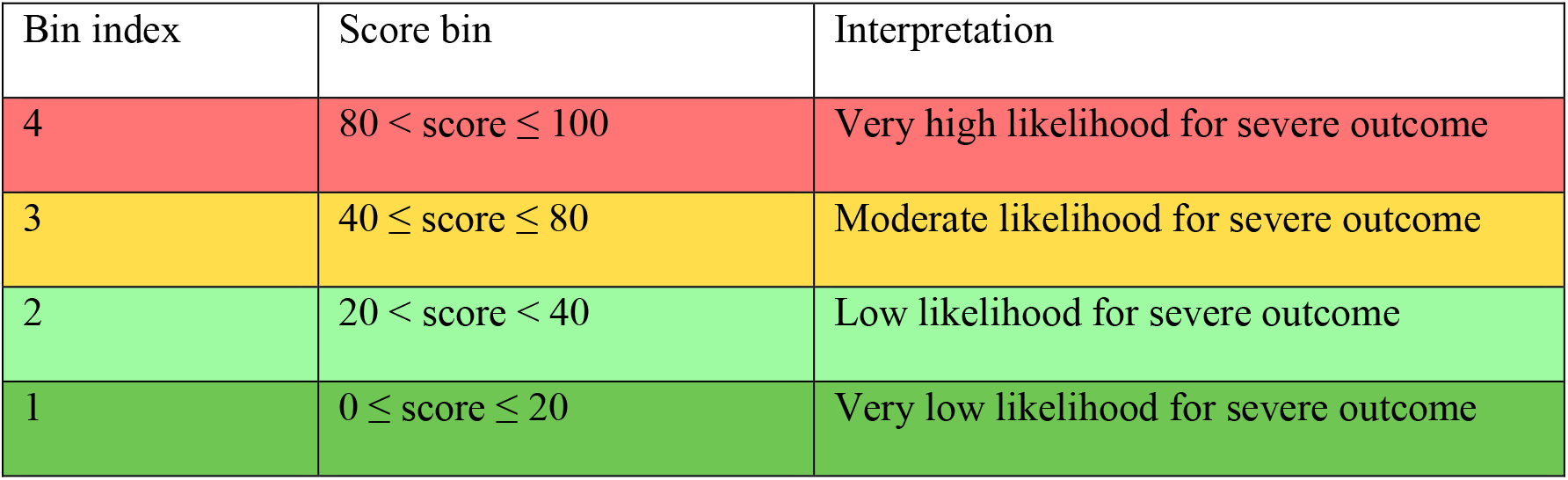
Score bin definition

### Prognostic value of the signature score

For a severity stratification tool to be predictive, its output must not only correlate with observable disease severity, but also identify those individuals who appear stable but have a high likelihood to deteriorate. To test the signature’s potential to predict future severe outcomes, the sub-group of patients who met a severe outcome for the first time after the day of blood draw (n=29) was identified. The time from blood draw to first outcome ranged between 1-13 days with a median of 4 days and the likelihood of future severe outcome increased significantly (p < 0.001) across the score bins (Supplementary Table 7). This trend was significant also with the definition of future severe outcome restricted to intubation with mechanical ventilation (Supplementary Table 8).

To further evaluate the signature’s predictive value, patients were stratified according to the time between blood draw and meeting the first severe outcome. Three groups of severe patients were defined: those meeting a first severe outcome on the day of blood draw (n=29), 1-3 days after (n=14), or more than 3 days after blood draw (n=15). Deterioration occurring in proximity to blood draw was associated with higher signature score; median scores were 87, 72, and 60 for the same-day, 1-3 days after, and >3 days after groups, respectively (Figure 5). Notably, the scores of each of the three severe patient groups, including the group meeting a first outcome over 3 days after blood draw, were significantly (p < 0.001) higher than the scores of patients who did not meet a severe outcome (n = 281).

**Figure 5.**
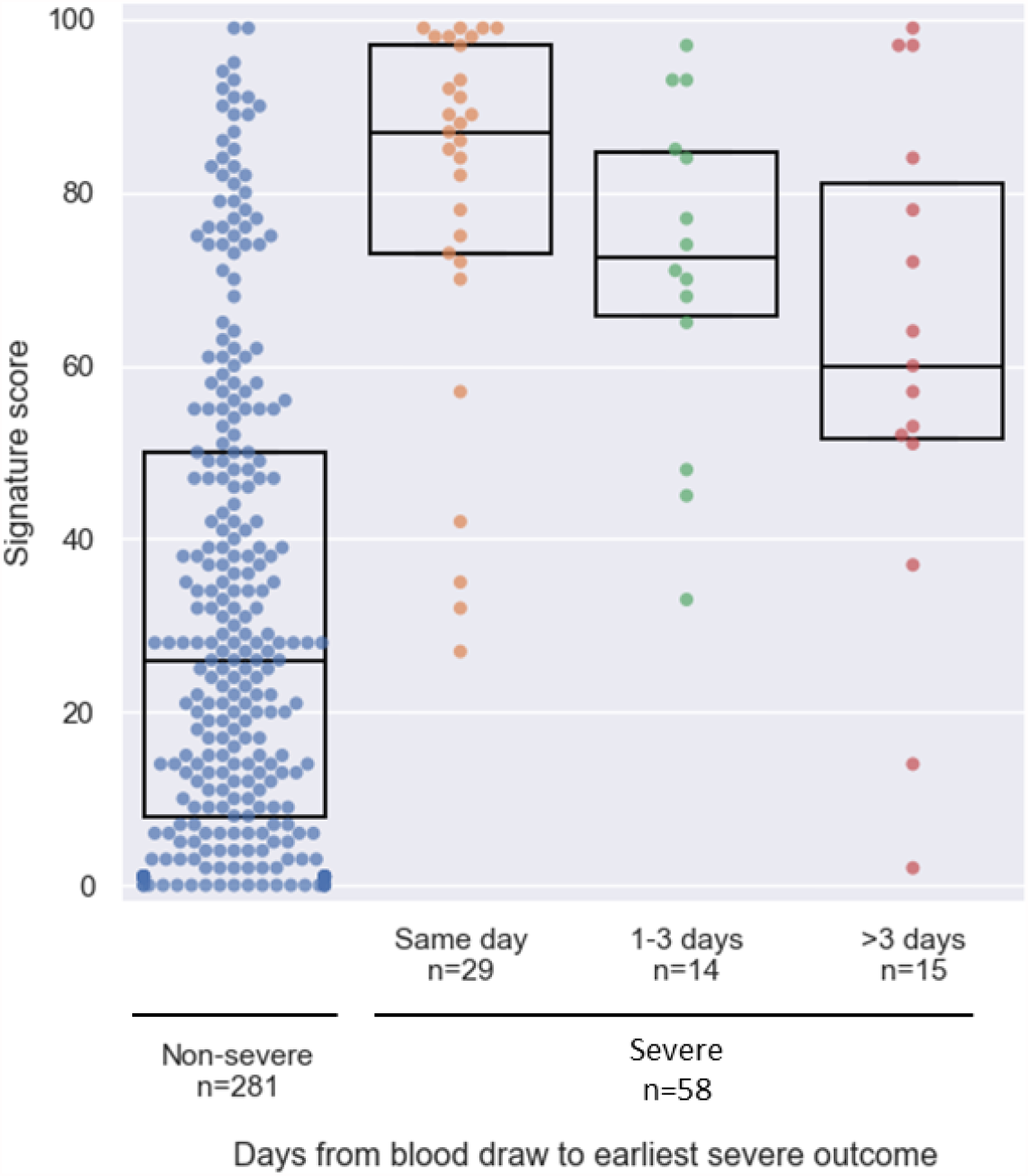
Signature score in severe patients meeting severity outcome on or after the day of blood draw stratified by time to first severe outcome, in comparison to non-severe patients (left). Dots represents patients and boxes denote median and IQR.

Finally, it was examined if the signature score provides added value beyond one of the parameters measured to assess the respiratory status of COVID-19 patients, namely SpO2 values. Six patients were identified with SpO2 > 93% on the day of blood draw who subsequently met a severe outcome, all of whom were assigned a signature score falling within the upper two bins (score range 60-85; Supplementary Table 9). This finding supports the signature’s potential to detect early-on patients who are at risk of future deterioration albeit with normal saturation (SpO2 > 93%) at time of blood draw.

### Signature’s potential to predict further deterioration and mortality

To evaluate the signature’s potential to predict further deterioration of severe patients, we identified the sub-group of severe patients who were admitted to the ICU and/or required non-invasive ventilation before blood draw (n=19). For this sub-group, deterioration was defined as requiring intubation with mechanical ventilation and/or death following the time of blood draw. Six out of the nineteen patients deteriorated, the time from blood draw to deterioration ranging between 0-13 days, with a median of 6.5 days. The remaining thirteen patients recovered, based on treatment termination and hospital discharge. The patients who deteriorated exhibited higher signature scores compared to those who recovered, with median score of 94.5 vs. 65 (Figure 6; p = 0.004). Lastly, the predictive value of the signature was assessed by examining survival prognosis. Thirty patients (8% of the cohort) died within 14 days of blood draw. 14-day survival probability was calculated using Kaplan-Meier estimator across the score bins. Survival distribution was significantly different comparing score bins 1+2 (score < 40) versus bins 3+4 (score ≥ 40) (Figure 7; p < 0.001).

**Figure 6.**
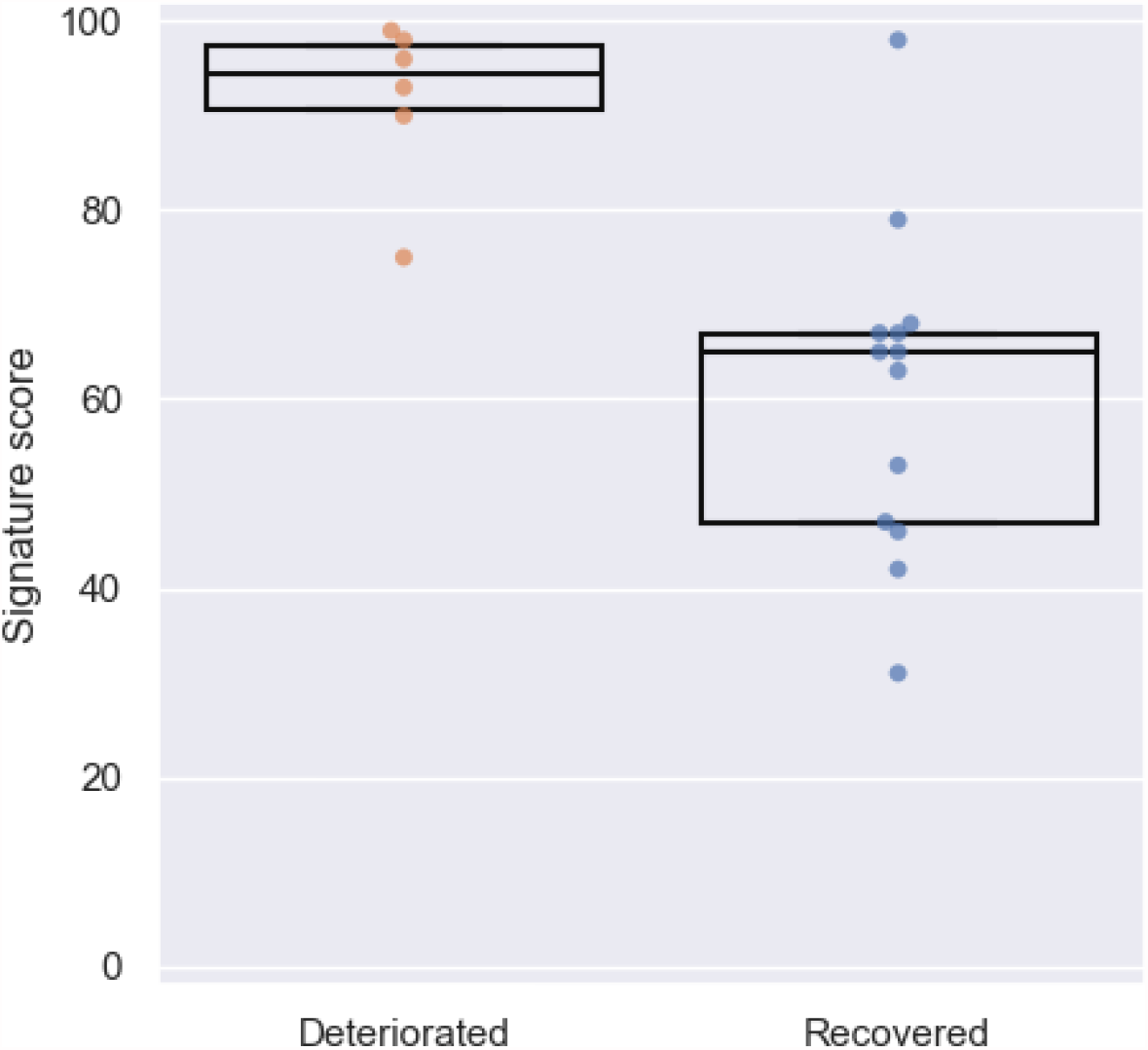
Score distribution in severe patients who further deteriorated (n=6) following blood draw versus those who recovered (n=13). Dots represent patients and boxes denote median and IQR.

**Figure 7.**
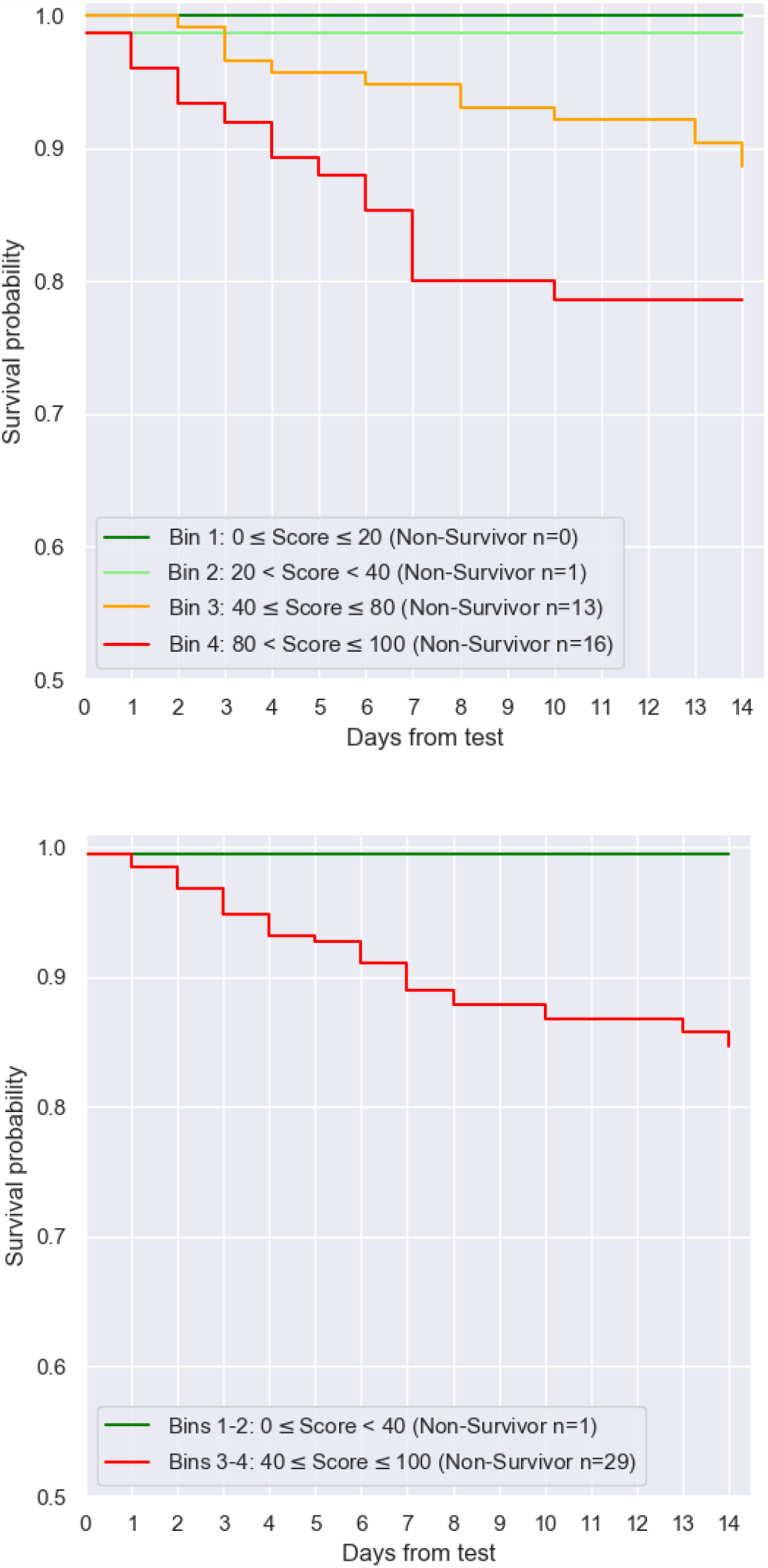
Kaplan-Meier survival estimates for signature score bins

## DISCUSSION

The present study describes derivation of a host-protein signature comprising TRAIL, IP-10 and CRP for accurately stratifying the severity of COVID-19 disease based on a multinational cohort recruited across multiple COVID-19 waves. The signature score was shown to accurately predict clinical deterioration among patients throughout the acute disease course. Patients with a severity score falling in the upper bins had higher likelihood to meet the composite severity outcome.

The host-protein signature demonstrated higher performance than other candidate severity tools examined in this study and performed irrespective of known risk factors and potential confounders, including age, sex, time from ED arrival, and comorbidities. Importantly, high performance was attained in the sub-cohort of patients meeting the severity outcome after the sampling day, supporting the signature’s prognostic value. Furthermore, the signature predicted deterioration even for patients that met severe outcomes more than three days after blood draw, supporting capability to detect severe outcomes early on. Patients that deteriorated in proximity to the day of blood draw exhibited higher scores than patients who deteriorated later. The trend in the score distribution across time suggests that higher scores may not only indicate the probability of severe outcomes but also give timing information. The tool’s prognostic value is further corroborated by two additional findings. First, among the patients who were admitted to the ICU or subjected to non-invasive ventilation before the day of blood draw, those that deteriorated further exhibited a significantly higher score. Second, in the sub-group of patients with normal saturation levels (SpO2 > 93%) the signature successfully assigned a high score to all patients who deteriorated following the day of blood testing. The latter result underscores the added clinical value of this tool beyond today’s standard of care in stratifying the severity of patients that appear similar according to their vital signs.

In addition to detecting early on who is likely to deteriorate, a severity tool can help to reduce unnecessary treatments or hospital admission for low-risk patients. In this regard, it is notable that almost 20% of cohort patients were hospitalized for 1-2 days and have not met any of the severe outcomes, potentially representing patients that may not have required admission. It has been recognized since the discovery of CRP as an inflammation marker (30), that host responses to infection have potential to serve as clinical decision-making tools. Advancements in host-response profiling and machine-learning algorithms have enabled a new generation of signatures to be developed, including one based on TRAIL, IP-10 and CRP for differentiating between bacterial and viral infection (23, 24, 39). Multiple studies have shown that these three immune proteins change expression in response to infection etiology (22–24, 31, 32) and also in response to the severity of infection (18–20, 27). The fact that these three proteins specifically change expression in the progression of COVID-19 disease (15, 25, 28, 29, 33–35, 40), prompted the present study to leverage a platform for their rapid measurement to derive a COVID-19 specific signature for severity stratification. It appears that immune dysregulation is pivotal to disease progression in SARS-CoV-2 infections and that IP-10 may be important in the development of lung damage, possibly when its expression is dissociated from interferon gamma levels (13, 25, 26). Of note, IP-10 and CRP have been shown to exhibit severity-related expression changes in SARS-CoV and middle east respiratory syndrome (MERS) infections, supporting that their response is not strain-specific but likely associated with viruses causing lung pathology (41, 42). In general, the window into the immune response to SARS-CoV-2 infection provided specifically by TRAIL, IP-10 and CRP, as individual biomarkers and as an integrative signature, has potential to provide clinicians with insight into disease course and patient management.

COVID-19 patients with severe or critical disease typically suffer respiratory failure and frequently require non-invasive or invasive respiratory support (https://www.covid19treatmentguidelines.nih.gov/). Accordingly, the rationale for the composite severity endpoint which guided signature development was to identify patients likely to exhibit respiratory insufficiency and require intense respiratory support. Treatment protocols have been dynamic during the pandemic and therefore multiple respiratory support methods were included (43, 44). Given that the multiple respiratory methods included in the composite endpoint represent varying levels of respiratory insufficiency, patients already meeting one of the severe outcomes before sampling were included in the derivation cohort, as these cases may deteriorate further and in a real world setting may benefit from a risk stratification tool. Indeed, this assumption was supported by our finding that severe patients who deteriorated further exhibited higher scores compared to severe patients who recovered. A major strength of the study is that the population was recruited across multiple international settings and multiple waves of the pandemic, supporting the generalizability of the findings. Broad applicability is also supported by similarity of the signature performance to cross-validation performance. Typically, studies that evaluate risk stratification tools for COVID-19 patient management focus on two severity outcomes, intubation with mechanical ventilation and/or mortality. Another strength of this study is the composite severity outcome, as prediction of requirement for non-invasive ventilation and ICU admission gives healthcare providers additional information that may not be obvious at patient evaluation, enabling better patient management.

An inherent limitation of this study is its retrospective design allowing only routinely measured data parameters to be evaluated. As a result, signature performance could not be compared to other commonly used severity scores or biomarkers and certain data points were not available (e.g., time from symptom onset) or missing. Also, the study period did not encompass multiple genetic variants of SARS-CoV-2. These limitations should be addressed in future independent signature validation studies. The signature was derived based on a population treated under protocols applied in the first and second waves of the pandemic, and its performance has not yet been evaluated in populations treated with newer approaches.

Future studies are required to validate the performance of this novel severity signature and to establish its utility in the evolving workflow of COVID-19 patient management, specifically in supporting decisions on escalation and de-escalation of care. A notable benefit of this immune-based tool, as compared to severity scores already in use, is that it predicts immune dysregulation associated with lung damage (25–27). Furthermore, unlike other severity scores, the three constituent proteins and signature can be easily and rapidly measured at the point-of-need using one platform. In summary, the derived signature together with the rapid measurement platform have potential to serve as a practical predictive tool for early indication of COVID-19 patients at-risk for severe outcome, facilitating improved patient management and outcomes.

## METHODS

### Study Design and Setting

This was an observational, convenience sampling study. Study cohort was composed of SARS-CoV-2-positive adult patients recruited retrospectively at 6 medical centers in the emergency department (ED), wards, and intensive care unit (ICU) between March and November 2020.

Inclusion criteria were: aged 18 years or older; SARS-CoV-2-positive reverse transcription polymerase chain reaction (RT-PCR) result; and a serum blood sample drawn within a week from ED arrival.

Patient data including past medical history, physical examination, laboratory data and medical treatment during hospitalization course were extracted from electronic medical records. Follow-up phone calls and verification of hospital re-admission status were performed to complete the 14-day observation period for patients discharged earlier. Alignment with the Transparent Reporting of a multivariable prediction model for Individual Prognosis Or Diagnosis (TRIPOD) Statement is indicated by responses given in the TRIPOD checklist (Supplementary Table 10) (37).

### Definition of clinical outcomes

Severe outcome was defined as mortality or respiratory failure requiring ICU admission, non-invasive ventilation (high flow nasal cannula, continuous positive airway pressure or bi-level airway pressure) or intubation with mechanical ventilation before or within 14 days from blood draw.

Levels of severe outcomes:

- level 1, ICU admission or non-invasive ventilation
- level 2, intubation with mechanical ventilation
- level 3, mortality

Among patients who met a severe outcome prior to blood draw, clinical deterioration was defined as transition from level 1 to level 2 or 3, and clinical recovery was defined as successful de-escalation of care (i.e., treatment termination and/or hospital discharge).

### Sample processing and measurement

At three medical centers (Hillel Yaffe, IL; Shaare Zedek, IL; Maimonides, USA), blood samples were processed to serum and frozen at -20ºC or - 80ºC and measured later. Blood samples from Shaare Zedek medical center were derived from the site’s biobank. At the other three medical centers (Hasharon, IL; Beilinson, IL; Saarland University, GER), blood samples were processed to serum and measured fresh.

Levels of TRAIL, IP-10, and CRP were measured using the MeMed Key® platform, which measures the concentrations of the three analytes in parallel from a single 100 µl serum sample using chemiluminescence technology within 15 minutes. The upper limit of quantitation was 300 pg/ml for TRAIL, 6,000 pg/ml for IP-10, and 250 mg/L for CRP. Procalcitonin assay was performed on the Roche Cobas e601 analyzer using the Elecsys® BRAHMS™ PCT kit (Roche Diagnostics, Switzerland). IL-6 assay was performed on the Roche Cobas e601 analyzer using the Elecsys® IL-6 kit (Roche Diagnostics, Switzerland). Other biomarkers levels such as ferritin, platelet count, and white blood cells were based on patient medical records.

### Missing data

Complete data for TRAIL, IP-10 and CRP predictor proteins were available for all cohort patients. A complete 14-day observation period was available for 96% of cohort patients; 15 patients (4%) discharged earlier and lost to follow-up were censored in calculation of Kaplan-Meier survival estimates, and treated as non-severe in the remaining analyses. Data on some of the clinical characteristics were not available for all 394 cohort patients (Supplementary Table 11); statistical analyses of these characteristics were based on available data.

The exact date of the non-invasive ventilation outcome was missing for 19 patients, and verified to occur before or within 14 days of blood draw based on the discharge date or medical record review. These patients were excluded from analyses stratifying patients according to time from blood draw to the first severe outcome.

### Multivariable model construction and score bin definition

The signature score is based on a model trained using L2-regularized logistic regression with balanced class weights. An additional transformation was applied to elevate the model-predicted probability as a non-linear function of CRP concentration. The resulting probability is rounded to an integer between 0-100.

Cross-validation was performed using logistic regression with the same configuration used to train the model, in a “leave-site-out” fashion: in each iteration all patients from one of the six medical centers were left out as a test set, and a model was trained based on patients from the other five sites and used to predict the probability of meeting a severe outcome for patients in the test set. Predicted probabilities from the six test sets were then combined into a single list for AUC calculation. Leave-site-out cross-validation evaluates the performance of a model in medical centers it was not trained on.

To generate a clinically intuitive tool for risk stratification, four score bins were defined. Each patient was assigned to a specific bin based on their signature score, and within the bin according to their severity outcome (Table 4).

The signature is called MeMed COVID-19 Severity™.

### Statistics

The distribution of variables was compared between two patient groups using two-sided Mann–Whitney *U* test for numeric variables, and two-sided Fisher’s exact test for binary variables. Two-sided statistical significance of the difference between ROC curves was calculated using the method of Hanley and McNeil (45). One-sided Cochran–Armitage (CA) test for trend was used to reject the null hypothesis that there is no trend of increasing probability of severe infection with higher score bin. The likelihood ratio (LR) of a score bin was calculated as the fraction of severe patients in that bin out of all severe patients, divided by the fraction of non-severe patients in the bin out of all non-severe patients.

Survival probabilities were calculated using Kaplan-Meier estimator. The statistical significance of the difference between the survival distribution of two groups was calculated with the log-rank test.

### Study approval

Ethical approval was obtained from each medical center with informed consent covered by the original protocol.

## Data Availability

The datasets generated during and/or analysed during the current study are available from the corresponding author on reasonable request.

## AUTHOR CONTRIBUTIONS

Niv Samuel Mastboim, Alon Angel, Oded Shaham, Tanya M Gottlieb, Eran Eden: Conceived and designed the study, analyzed data (including signature derivation) and wrote the manuscript.

Tahel Ilan Ber, Eran Reiner: Designed the study, analyzed data and reviewed the manuscript. Roy Navon, Paul Feigin: Analyzed data (including signature derivation) and reviewed the manuscript.

Einav Simon, Michal Rosenberg, Yael Israeli, Mary Hainrichson, Noa Avni: Conducted laboratory measurements, analyzed data and reviewed the manuscript.

Kfir Oved: Conceived the study and reviewed the manuscript.

Boaz Tadmor, Pierre Singer, Ilya Kagan, Shaul Lev, Daniel Diker, Amir Jarjou’i, Ramzi Kurd, Eli Ben-Chetrit, Guy Danziger, Cihan Papan, Sergey Motov, Ma’anit Shapira, Michal Stein, Adi Klein: Enrolled patients and reviewed the manuscript.

## ACKNOWLEDGEMENTS

We thank Dr Oren Zarchin, Dr Moran Barak and Dr Efrat Hartog-David for their professional insights. We thank Yaly Orr, Idan Lancry, Dvir Ilan, Noa Kremer, Ruth Yudalevich, Adi Cohen, Michal Largman and Dennis Kuhn for their dedicated data management and Noa Serruya for laboratory support. We thank Einat Moscoviz for leading the clinical coordination.

