## Supplemental materials for "An immune-protein signature combining TRAIL, IP-10 and CRP for accurate prediction of severe COVID-19 outcome"

Supplementary Materials

| *Medical center* | *Maimonides* | *Saarland University Hospital* | *Hillel Yaffe* | *Shaare Zedek* | *Beilinson* | *Hasharon* |
|---|---|---|---|---|---|---|
| *City, Country* | New York, US | Homburg, GER | Hadera, ISR | Jerusalem, ISR | Petah Tikva, ISR | Petah Tikva, ISR |
| *Number of beds* | 711 | 1202 | 515 | 814 | 835 | 241 |
| *Enrollment period* | July-Nov 2020 | Mar-Apr 2020 | Mar-Oct 2020 | Apr-Aug 2020 | July-Nov 2020 | Apr-May 2020 |
| *Enrollment setting* | ED, Ward, ICU | Ward, ICU | ED | ED, Ward, ICU | Wards, ICU | Ward, ICU |
| *N (Severe/Non severe)* | 42 (18/24) | 13 (9/4) | 103 (18/85) | 119 (30/89) | 74 (30/44) | 43 (8/35) |

Supplementary Table 1: Medical centers included in derivation study.

| Assay score | Age (range) | Sex | Time from ED admission to blood sample (days) | Time from first COVID-19 confirmation to blood sample (days) | Time from symptom onset to blood sample (days) | Severe outcome met | Time from blood sample to first severe outcome (days) | Immuno-modulation drugs prior to blood sample | Duration of Immuno-modulation treatment (days) | Medications given during hospitalization | Hospital course |
|---|---|---|---|---|---|---|---|---|---|---|---|
| 2 | 36-40 | F | 0 | 3 | 5 | High flow nasal canula | 4 | Prednisone | 1 | Convalescence plasma, Remdesivir, Ceftriaxone, Azithromycin, Dexamethasone | * Five days history of flu like symptoms<br>* Physical examination and chest X-ray indicative of COVID-19 pneumonia<br>* Treated with low flow oxygen canula and was later put on high flow.<br>* Total length of stay of 10 days |
| 8 | 61-65 | M | 6 | 8 | 16 | ICU, Mechanical ventilation | -6 (Intubated prior to blood sample) | Actemra | 3 | Ceftriaxone, Azithromycin, Hydroxychloroquine Solumedrol | * Ten days history of fever, cough, dyspnea.<br>* intubated and transferred to the ICU on day of admission.<br>* Study blood sample collected 6 days after ICU admission, while presenting signs of clinical improvement<br>* Extubated 5 days following study sample collection<br>* Total length of stay of 17 days. |
| 14 | 86-90 | F | 0 | 6 | 6 | High flow nasal canula | 9 | Unknown | Unknown | None | * Five days history of desaturation measured at nursing home.<br>* No abnormal findings on chest-x-ray or physical examination<br>* Treated with low flow nasal canula and was put on high flow after 9 days.<br>* Total length of stay of 14 days. |

Supplementary Table 2: Clinical data of patients who met a severe outcome and were assigned to the lowest bin (false negatives).

| Bin | Score | % Patients | n Total | n Death/ IMV | n No Death/ IMV | % Death/ IMV (PPV) | % No Death/ IMV (NPV) | LR (95% CI) |
|---|---|---|---|---|---|---|---|---|
| 4 | 80 < score ≤ 100 | 19.04 | 75 | 34 | 41 | 45.33 | 54.67 | 4.53 (3.15-6.51) |
| 3 | 40 ≤ score ≤ 80 | 29.44 | 116 | 23 | 93 | 19.83 | 80.17 | 1.35 (0.94-1.95) |
| 2 | 20 < score < 40 | 20.05 | 79 | 3 | 76 | 3.80 | 96.20 | 0.22 (0.07-0.66) |
| 1 | 0 ≤ score ≤ 20 | 31.47 | 124 | 1 | 123 | 0.81 | 99.19 | 0.04 (0.01-0.31) |
| | **Total** | 100.00 | 394 | 61 | 333 | | | |

Supplementary Table 3: Distribution of patients across score bins according to severe outcome, with the definition of severe outcome limited to intubation with mechanical ventilation or death.

IMV, intubation with mechanical ventilation. LR, likelihood ratio. PPV, positive predictive value. NPV, negative predictive value.

| Bin | Score | Male n | Male n Severe | Male % severe | Male LR (95%CI) | Female n | Female n Severe | Female % severe | Female LR (95%CI) |
|---|---|---|---|---|---|---|---|---|---|
| ***Sex*** | | | | | | | | | |
| *4* | **80 < score ≤ 100** | 54 | 41 | 75.93 | 6.03 (3.44-10.58) | 21 | 13 | 61.90 | 6.30 (2.85-13.93) |
| *2* | **40 ≤ score ≤ 80** | 74 | 32 | 43.24 | 1.46 (1.00-2.11) | 42 | 14 | 33.33 | 1.94 (1.16-3.25) |
| *2* | **20 < score < 40** | 51 | 6 | 11.76 | 0.26 (0.11-0.57) | 28 | 4 | 14.29 | 0.65 (0.24-1.73) |
| *1* | **0 ≤ score ≤ 20** | 54 | 1 | 1.85 | 0.04 (0.01-0.26) | 70 | 2 | 2.86 | 0.11 (0.03-0.44) |
| | **Total** | 233 | 80 | | | 161 | 33 | | |

| Bin | Score | 65 years or younger n | n Severe | % severe | LR (95%CI) | Over 65 years n | n Severe | % severe | LR (95%CI) |
|---|---|---|---|---|---|---|---|---|---|
| ***Age*** | | | | | | | | | |
| *4* | **80 < score ≤ 100** | 36 | 26 | 72.22 | 7.80 (4.01-15.16) | 39 | 28 | 71.79 | 5.00 (2.69-9.28) |
| *2* | **40 ≤ score ≤ 80** | 56 | 22 | 39.29 | 1.94 (1.24-3.03) | 60 | 24 | 40.00 | 1.31 (0.87-1.96) |
| *2* | **20 < score < 40** | 45 | 7 | 15.56 | 0.55 (0.26-1.17) | 34 | 3 | 8.82 | 0.19 (0.06-0.59) |
| *1* | **0 ≤ score ≤ 20** | 91 | 2 | 2.20 | 0.07 (0.02-0.27) | 33 | 1 | 3.03 | 0.06 (0.01-0.44) |
| | **Total** | 228 | 57 | | | 166 | 56 | | |

| Bin | Score | Two days or less n | n Severe | % severe | LR (95%CI) | Over two days n | n Severe | % severe | LR (95%CI) |
|---|---|---|---|---|---|---|---|---|---|
| ***Time from ED arrival to blood draw*** | | | | | | | | | |
| *4* | **80 < score ≤ 100** | 53 | 37 | 69.81 | 7.09 (4.18-12.01) | 22 | 17 | 77.27 | 4.08 (1.67-9.94) |
| *2* | **40 ≤ score ≤ 80** | 87 | 31 | 35.63 | 1.70 (1.19-2.42) | 29 | 15 | 51.72 | 1.29 (0.72-2.28) |
| *2* | **20 < score < 40** | 69 | 8 | 11.59 | 0.40 (0.20-0.80) | 10 | 2 | 20.00 | 0.30 (0.07-1.32) |
| *1* | **0 ≤ score ≤ 20** | 108 | 2 | 1.85 | 0.06 (0.01-0.23) | 16 | 1 | 6.25 | 0.08 (0.01-0.58) |
| | **Total** | 317 | 78 | | | 77 | 35 | | |

| Bin | Score | Less than two n | n Severe | % severe | LR (95%CI) | Two or more n | n Severe | % severe | LR (95%CI) |
|---|---|---|---|---|---|---|---|---|---|
| ***Number of Comorbidities***[A] | | | | | | | | | |
| *4* | **80 < score ≤ 100** | 52 | 36 | 69.23 | 6.40 (3.77-10.87) | 23 | 18 | 78.26 | 6.07 (2.47-14.90) |
| *2* | **40 ≤ score ≤ 80** | 82 | 34 | 41.46 | 2.02 (1.41-2.88) | 34 | 12 | 35.29 | 0.92 (0.52-1.62) |
| *2* | **20 < score < 40** | 62 | 6 | 9.68 | 0.30 (0.14-0.68) | 17 | 4 | 23.53 | 0.52 (0.18-1.47) |
| *1* | **0 ≤ score ≤ 20** | 104 | 2 | 1.92 | 0.06 (0.01-0.22) | 20 | 1 | 5.00 | 0.09 (0.01-0.63) |
| | **Total** | 300 | 78 | | | 94 | 35 | | |

Supplementary Table 4: Signature performance in sub-populations.
[A]The comorbidities considered were hypertension, diabetes mellitus, chronic heart failure, and malignancy (Table 1).

| Added Feature | AUC (95% CI) |
|---|---|
| N/A | 0.86 (0.81-0.90) |
| Sex | 0.85 (0.81-0.90) |
| Age | 0.85 (0.80-0.90) |
| Time from ED arrival to blood draw (days) | 0.86 (0.81-0.90) |
| Number of comorbidities[A] | 0.85 (0.81-0.90) |

Supplementary Table 5: Contribution of additional parameters to cross-validation performance. AUC, area under the receiver operating characteristic curve. CI, confidence interval.
[A]The comorbidities considered were hypertension, diabetes mellitus, chronic heart failure, and malignancy (Table 1).

| | Total available | n Death/ IMV | n No Death/ IMV | Comparator AUC (95% CI) | Signature AUC (95% CI) | p-value |
|---|---|---|---|---|---|---|
| **Signature** | 394 | 61 | 333 | N/A | 0.85 (0.79-0.92) | N/A |
| **Age** | 394 | 61 | 333 | 0.69 (0.61-0.77) | 0.85 (0.79-0.92) | 0.001 |
| **Oxygen saturation** | 251 | 24 | 227 | 0.65 (0.52-0.77) | 0.88 (0.79-0.97) | 0.001 |
| **CRP** | 394 | 61 | 333 | 0.79 (0.72-0.86) | 0.85 (0.79-0.92) | 0.013 |
| **IL-6** | 139 | 13 | 126 | 0.78 (0.63-0.93) | 0.87 (0.74-0.99) | 0.305 |
| **PCT** | 106 | 11 | 95 | 0.75 (0.58-0.92) | 0.87 (0.73-1.00) | 0.209 |
| **Ferritin** | 71 | 11 | 60 | 0.63 (0.44-0.82) | 0.75 (0.58-0.93) | 0.257 |

Supplementary Table 6: Comparison of the signature performance to other parameters and biomarkers, with the definition of severe outcome limited to intubation with mechanical ventilation or death. The analysis was done on the subset of patients with available measurements for each comparator. IMV, intubation with mechanical ventilation. AUC, area under the receiver operating characteristic curve. CI, confidence interval.

| Bin | Score | % Patients | n Total | n Severe | n Non-Severe | % Severe (PPV) | % Non-Severe (NPV) | LR (95% CI) |
|---|---|---|---|---|---|---|---|---|
| **4** | **80 < score ≤ 100** | 9.68 | 30 | 9 | 21 | 30.00 | 70.00 | 4.15 (2.10-8.20) |
| **3** | **40 ≤ score ≤ 80** | 27.74 | 86 | 16 | 70 | 18.60 | 81.40 | 2.21 (1.51-3.26) |
| **2** | **20 < score < 40** | 22.90 | 71 | 2 | 69 | 2.82 | 97.18 | 0.28 (0.07-1.09) |
| **1** | **0 ≤ score ≤ 20** | 39.68 | 123 | 2 | 121 | 1.63 | 98.37 | 0.16 (0.04-0.61) |
| | **Total** | 100.00 | 310 | 29 | 281 | | | |

Supplementary Table 7: Distribution of patients across score bins, with severe patients restricted to those who met a severe outcome for the first time after the day of blood draw.

LR, likelihood ratio. PPV, positive predictive value. NPV, negative predictive value.

| Bin | Score | % Patients | n Total | n IMV | n Non-IMV | % IMV (PPV) | % Non-IMV (NPV) | LR (95% CI) |
|---|---|---|---|---|---|---|---|---|
| **4** | **80 < score ≤ 100** | 15.26 | 56 | 11 | 45 | 19.64 | 80.36 | 3.83 (2.33-6.31) |
| **3** | **40 ≤ score ≤ 80** | 29.97 | 110 | 9 | 101 | 8.18 | 91.82 | 1.40 (0.82-2.37) |
| **2** | **20 < score < 40** | 21.25 | 78 | 2 | 76 | 2.56 | 97.44 | 0.41 (0.11-1.57) |
| **1** | **0 ≤ score ≤ 20** | 33.51 | 123 | 0 | 123 | 0.00 | 100.00 | 0.00 |
| | **Total** | 100.00 | 367 | 22 | 345 | | | |

Supplementary Table 8: Distribution of patients across score bins, with severe outcome defined as intubation with mechanical ventilation (IMV) after the day of blood draw. Patients who had met the IMV outcome on the day of blood draw (n = 9) or before (n = 18) were excluded. Of note, applying a single threshold of 40 score units across the entire cohort (n = 367) yields sensitivity of 91%, specificity of 58%, negative predictive value of 99% and positive predictive value of 12%.

LR, likelihood ratio. PPV, positive predictive value. NPV, negative predictive value

| | *Non-severe patients (n)* | | *Severe patients (n)* | | |
|---|---|---|---|---|---|
| | *SpO2>93%* | *SpO2≤93%* | *SpO2>93%* | *SpO2≤93%* | Total |
| *Bin 1* | 75 | 15 | 0 | 1 | 91 |
| *Bin 2* | 33 | 21 | 0 | 2 | 56 |
| *Bin 3* | 20 | 17 | 4 | 4 | 45 |
| *Bin 4* | 5 | 6 | 2 | 3 | 16 |
| *Total* | 133 | 59 | 6 | 10 | 208 |

Supplementary Table 9: Distribution of saturated (SpO2>93%) vs. desaturated (SpO2≤93% patients across signature score bins. SpO2 was measured on the day of blood draw. Severe patients were restricted to those meeting a severe outcome for the first time after the day of blood draw. SpO2, Oxygen saturation on room air.
SpO2 cutoff of 93% was used in line with NIH classification of disease severity (https://www.covid19treatmentguidelines.nih.gov/).

| Section/Topic | | Checklist Item | Status |
|---|---|---|---|
| **Title and abstract** | | | |
| Title | 1 | Identify the study as developing and/or validating a multivariable prediction model, the target population, and the outcome to be predicted. | Done |
| Abstract | 2 | Provide a summary of objectives, study design, setting, participants, sample size, predictors, outcome, statistical analysis, results, and conclusions. | Done |
| **Introduction** | | | |
| Background and objectives | 3a | Explain the medical context (including whether diagnostic or prognostic) and rationale for developing or validating the multivariable prediction model, including references to existing models. | Done |
| | 3b | Specify the objectives, including whether the study describes the development or validation of the model or both. | Done |
| **Methods** | | | |
| Source of data | 4a | Describe the study design or source of data (e.g., randomized trial, cohort, or registry data), separately for the development and validation data sets, if applicable. | Done |
| | 4b | Specify the key study dates, including start of accrual; end of accrual; and, if applicable, end of follow-up. | Done |
| Participants | 5a | Specify key elements of the study setting (e.g., primary care, secondary care, general population) including number and location of centres. | Done |
| | 5b | Describe eligibility criteria for participants. | Done |
| | 5c | Give details of treatments received, if relevant. | Done |
| Outcome | 6a | Clearly define the outcome that is predicted by the prediction model, including how and when assessed. | Done |
| | 6b | Report any actions to blind assessment of the outcome to be predicted. | |
| Predictors | 7a | Clearly define all predictors used in developing or validating the multivariable prediction model, including how and when they were measured. | Done |
| | 7b | Report any actions to blind assessment of predictors for the outcome and other predictors. | |
| Sample size | 8 | Explain how the study size was arrived at. | Done |
| Missing data | 9 | Describe how missing data were handled (e.g., complete-case analysis, single imputation, multiple imputation) with details of any imputation method. | Done |
| Statistical analysis methods | 10a | Describe how predictors were handled in the analyses. | |
| | 10b | Specify type of model, all model-building procedures (including any predictor selection), and method for internal validation. | Done |
| | 10d | Specify all measures used to assess model performance and, if relevant, to compare multiple models. | |
| Risk groups | 11 | Provide details on how risk groups were created, if done. | |
| **Results** | | | |
| Participants | 13a | Describe the flow of participants through the study, including the number of participants with and without the outcome and, if applicable, a summary of the follow-up time. A diagram may be helpful. | Done |
| | 13b | Describe the characteristics of the participants (basic demographics, clinical features, available predictors), including the number of participants with missing data for predictors and outcome. | Done |
| Model development | 14a | Specify the number of participants and outcome events in each analysis. | Done |
| | 14b | If done, report the unadjusted association between each candidate predictor and outcome. | Done |
| Model specification | 15a | Present the full prediction model to allow predictions for individuals (i.e., all regression coefficients, and model intercept or baseline survival at a given time point). | |
| | 15b | Explain how to the use the prediction model. | Done |
| Model performance | 16 | Report performance measures (with CIs) for the prediction model. | Done |
| **Discussion** | | | |
| Limitations | 18 | Discuss any limitations of the study (such as nonrepresentative sample, few events per predictor, missing data). | Done |
| Interpretation | 19b | Give an overall interpretation of the results, considering objectives, limitations, and results from similar studies, and other relevant evidence. | Done |
| Implications | 20 | Discuss the potential clinical use of the model and implications for future research. | Done |
| **Other information** | | | |
| Supplementary information | 21 | Provide information about the availability of supplementary resources, such as study protocol, Web calculator, and data sets. | |
| Funding | 22 | Give the source of funding and the role of the funders for the present study. | |

Supplementary Table 10: TRIPOD checklist for Prediction Model Development.

| Parameter name | Total n | Severe n | Non-severe n |
|---|---|---|---|
| Length of hospital stay (days) | 382 | 106 | 276 |
| Heart rate (beats per minute) | 294 | 87 | 207 |
| Temperature (C˚) | 248 | 67 | 181 |
| Systolic blood pressure (mmHg) | 269 | 80 | 189 |
| Respiratory rate (breath/minute) | 108 | 45 | 63 |
| Oxygen saturation on room air (SpO2) | 251 | 59 | 192 |
| WBC (K/ml$^3$) | 158 | 43 | 115 |
| Ferritin (ng/ml) | 71 | 31 | 40 |
| PCT (ng/ml) | 106 | 29 | 77 |
| IL-6 (pg/ml) | 139 | 33 | 106 |

Supplementary Table 11: Parameters for which data was available for a subset of the 394 cohort patients.

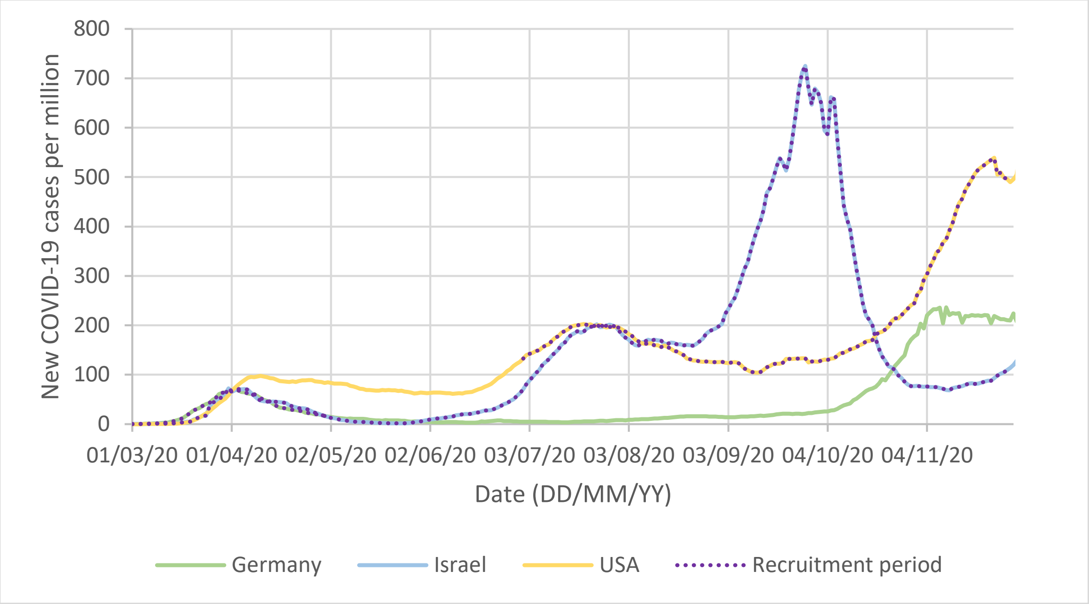


Supplementary Figure 1: Recruitment period encompassed first and second COVID-19 waves. Number of new SARS-CoV-2-positive cases detected by PCR per day per million individuals was sourced from The Johns Hopkins University dashboard and dataset[51]
